# Plasma Proteomics for Parkinson’s Disease: Diagnostic Classification, Severity Association, and Therapeutic Hypotheses

**DOI:** 10.1101/2025.09.02.25334526

**Authors:** Nicholas Minster, M. Saleet Jafri

## Abstract

**Background:** Although there is no cure, early diagnosis of Parkinson’s disease allows effective management and symptomatic relief, potentially delaying the need for more potent medications. Blood-based biomarkers can facilitate early detection, symptom monitoring, and targeted therapy.

**Methods:** Gene expression and plasma proteomics data from two Parkinson’s disease cohorts were integrated. Machine learning models were trained to classify disease status using protein, gene expression, and combined datasets. A severity score was derived by regressing protein levels against clinical motor ratings and tested longitudinally. Enrichment and network analyses identified biological context, and drug perturbation databases were queried for candidate therapies.

**Results:** Proteomic models outperformed gene-based approaches and generalized well to external data. The severity score correlated with clinical burden and predicted future progression. Enriched pathways involved extracellular signaling, immune response, and post-translational regulation. Several compounds were identified as potential therapeutic candidates based on network targeting and reversal potential.

**Conclusions:** Peripheral proteomic signatures offer classification, progression, and therapeutic insights in Parkinson’s disease, supporting their biological relevance.

## 1 Background

Parkinson’s disease (PD) represents a looming public health crisis, distinguished not only by its rising global burden but also by the persistent gap between biological onset and symptom onset. Parkinson’s Disease is now recognized as the fastest-growing neurodegenerative disorder. Diagnosis relies on motor signs that appear after substantial neurodegeneration has occurred [1, 2]. This delay limits opportunities for disease-modifying therapy. At the molecular level, PD is characterized by the progressive loss of dopaminergic neurons in the substantia nigra pars compacta and the pathological aggregation of *α*-synuclein into Lewy bodies [3]. Closing the diagnostic gap is a central challenge in Parkinson’s disease research [4].

Diagnosis of Parkinson’s disease (PD) relies on taking a patient history and neurological exam [5, 6]. Despite active biomarker research, available tests are limited in routine care. Advanced imaging such as DaTScan or high-field MRI is constrained by access and cost [7]. Emerging assays such as CSF *α*-synuclein seeding remain confined to research settings and are suitable only as a confirmation test [8, 9].

Monogenic forms of PD, although relatively rare, provide critical insight into the molecular pathways disrupted in both familial and sporadic cases. Mutations in GBA1 and LRRK2 together account for approximately 10–15% of cases and contribute to PD pathology through distinct yet intersecting mechanisms [10]. GBA1 mutations compromise lysosomal glucocerebrosidase activity, impairing autophagic clearance and promoting *α*-synuclein accumulation [11]. LRRK2 gain-of-function mutations alter vesicular trafficking and mitochondrial dynamics via dysregulated kinase signaling [12]. These disruptions converge on lysosomal-autophagic dysfunction and mitochondrial stress, hallmarks increasingly recognized across genetic and idiopathic PD [13]. Studying these mutations has revealed broader disease mechanisms and shared molecular targets for early diagnosis and intervention.

Within this landscape, the circulating secretome, the set of proteins and molecules secreted into the bloodstream, has emerged as a promising biomarker source. It reflects cellular stress, neuroinflammation, and systemic signaling changes that may occur before clinical symptoms arise.

Current therapies are palliative [14]. Levodopa and adjunctive dopaminergic agents alleviate symptoms but fail to halt disease progression and often lead to long-term motor complications [15, 16]. Invasive interventions such as deep brain stimulation offer motor symptom relief in select patients [17]. Multiple recent trials targeting *α*-synuclein aggregation and neuroinflammation have failed to demonstrate disease-modifying effects, underscoring the need for upstream biomarkers and therapeutic targets [18].

In parallel, lifestyle interventions such as physical exercise are gaining recognition as low-risk strategies that may confer neuroprotective benefits. Exercise has been shown to modulate DNA methylation patterns associated with aging and neurodegeneration, potentially reversing aspects of the epigenetic clock [19]. Exercise also enhances neuroplasticity, reduces neuroinflammation, and engages molecular pathways related to autophagy, mitochondrial function, and immune regulation, processes central to PD pathogenesis. These molecular effects position physical activity not only as a therapeutic adjunct but also as a window into modifiable disease mechanisms that precede clinical onset.

High-throughput transcriptomic and proteomic platforms enable comprehensive profiling of RNA and protein species, including secreted factors, from small plasma volumes. When paired with machine learning and network biology, these datasets can yield high diagnostic accuracy and identify molecular patterns predictive of severity and progression [20]. However, many studies lack rigorous cross-validation or external replication, raising concerns about generalizability [21, 22]. Interpretable machine learning tools like SHapley Additive exPlanations (SHAP) and Integrated Gradients offer a path forward by linking predictive features to known biology. Integrative multi-omics modeling may further enable the capture of complex molecular phenotypes [23]. These steps translate statistical signals into actionable biological insight by deriving blood-based biomarkers for (i) diagnostic classification (PD vs healthy control), (ii) severity monitoring via a Proteomic Severity Index (PSI), and (iii) hypothesis-generating targets to prioritize therapeutic mechanisms for follow-up.

### Study framing and departures from prior work

This work builds directly on the NIA-led study by Makarious and colleagues, which trained multimodal machine-learning models on PPMI and externally validated in PDBP [24]. In that foundation study, combining modalities (e.g., olfaction and polygenic risk alongside molecular data) outperformed single-modality approaches and achieved strong external AUCs. The effort was supported by the Intramural Research Program of the National Institute on Aging and NINDS—underscoring NIH’s strategic interest in predictive modeling for PD [24]. Predictors in this study were restricted to molecular features to quantify the stand-alone diagnostic and severity-tracking signal in peripheral molecules. This avoids clinical scales that can dominate models and dilute the biological signal.

To ensure defensible estimates and avoid information leakage, all preprocessing and feature selection are nested within participant-grouped cross-validation, with a prespecified internal hold-out and external testing only once model selection is complete. Early detection is a central motivation: accurate identification during the prodromal phase could enable earlier intervention and better trial enrichment [25]. PPMI is well-suited as a training resource for this aim, as the study explicitly recruits individuals with early, untreated (de novo) Parkinson’s disease and healthy controls of similar age and sex; PD eligibility required a positive dopamine transporter (DAT) SPECT scan on central read, and healthy controls showed normal DAT imaging at baseline.

## Methods

Matched whole-blood RNA sequencing and targeted plasma Olink proteomics data were obtained from the PPMI IR3 and 2023 PDBP releases, available through the AMP-PD Knowledge Platform [26]. These datasets were chosen because they represent two of the largest and most comprehensive longitudinal cohorts for Parkinson’s disease, offering deeply integrated clinical and multi-omic data for well-characterized patients. After excluding samples that failed internal quality control, the integrated dataset comprised 971 visit-level samples from 413 unique participants, including 58,294 genes and 1,463 proteins. For cross-sectional modeling, participants were required to have at least one longitudinal instance with RNA-seq, proteomics, and clinical (MDS-UPDRS) data. This criterion yielded 235 participants and 745 visit-level samples for longitudinal analyses (median = 3 visits, IQR = 2–4). Among these, 111 participants contributed a baseline (visit-0) draw with matched MDS-UPDRS scores.

The Olink dataset spanned neurology, inflammation, cardiometabolic, and oncology panels, with protein features identified by UniProt IDs. Only participants with both RNA-seq and proteomics data were retained for integrative analyses. For RNA-seq, differential expression analysis was performed using independent t-tests with Benjamini–Hochberg correction to reduce the number of features. The PPMI cohort comprised three groups: (1) Parkinson’s disease (PD) patients diagnosed within two years and not on medication, (2) prodromal individuals over 60 with PD risk factors, and (3) healthy controls without neurological disease or a family history of PD [27]. To ensure diagnostic clarity, participants from the SWEDD and Genetic Registry cohorts were excluded. For external validation, PDBP RNA-seq and proteomics data harmonized to PPMI protocols were incorporated [28].

RNA-seq and Olink proteomic features were concatenated as predictors of binary disease status (Parkinson’s disease versus healthy control), with the class label assigned accordingly for each sample. Within PPMI, 20% of PD samples were held out (stratified by patient ID) for internal testing, and all PDBP samples were reserved for external validation. To correct for class imbalance, the Random Forest classifier was configured with *class*_*w*_*eight* =^*′*^ *balanced*^*′*^. Random Forest was compared with candidate classifiers (XGBoost, Gradient Boosting, SVM, Logistic Regression, Extra Trees), and was selected based on the highest ROC-AUC. In each cross-validation fold (10-fold GroupKFold on participant ID), variance thresholding, SelectKBest (ANOVA F-statistic), and standard scaling were applied prior to classification. Random Forest hyperparameters were tuned using 50 Latin Hypercube–sampled parameter sets in RandomizedSearchCV, optimizing for ROC-AUC. Latin Hypercube sampling was chosen for its balance of efficiency and reproducibility, offering uniform coverage of the hyperparameter space while reducing the number of required parameter sets compared to grid or random search [29].

### 2.1 Analysis of Selected Features

Model explanations were generated with SHapley Additive exPlanations (SHAP; Python package shap v0.44.1), quantifying each feature’s contribution to PD-versus-control predictions [30]. SHAP values were computed relative to control class probability; negative values indicated features that increased PD probability. SHAP summary plots provided global and local interpretability, while waterfall plots visualized probability shifts for individual cases. Force plots highlighted correctly and incorrectly classified samples.

### 2.2 Gene Ontology Enrichment

Gene Ontology (GO) enrichment analysis was performed with the `g:Profiler` Python API, querying UniProt IDs against GO:BP, GO:MF, and GO:CC terms. Significant terms (*p <* 0.05) were identified, gene ratios were calculated as the proportion of input proteins mapping to each term, and exported ranked results as CSV files for downstream analysis.

### 2.3 PPI Network Construction

Protein–protein interaction (PPI) networks were constructed by querying STRING DB v11.5 (accessed January 2025) with the `STRINGdb` R package (v2.8.5). The search was restricted to *Homo sapiens*, required experimental or curated evidence, set the confidence threshold to 0.4, and excluded self-loops and disconnected nodes. Cytoscape 3.10.0 using the STRING App 2.0 was used to visualize the network.

### 2.4 Hub Identification

Network hubs were identified using the CytoNCA plugin in Cytoscape. Nodes with a degree *z*-score ≥ 2 were defined as hubs. Hub robustness was verified through sensitivity analyses using betweenness and closeness centrality.

### 2.5 Connectivity Map Query

Differentially regulated proteins (| log_2_ FC | ≥ 0.5, FDR *<* 0.05) were mapped to coding genes and queried the CLUE platform (CMap build 02) via the `cmapR` Python package (v4.0.2). Compounds with connectivity scores (tau) ≤ ± 90 were retained as candidate perturbagens.

### 2.6 Perturbagen Target Retrieval

Validated human targets were retrieved for each compound from DrugBank 5.3 (downloaded January 2025) and ChEMBL 32 via the `bioservices` Python API. Targets were collapsed to unique gene symbols and mapped onto the STRING PPI network.

### 2.7 Target Overlay and Enrichment Testing

Drug-target genes were overlaid on the PPI network using the enhancedGraphics Cytoscape plugin, representing compound–target edges as dashed lines [31]. Enrichment was assessed with one-sided Fisher’s exact tests, comparing observed target hits among PD-associated nodes to expectations across all ∼ 19,000 STRING proteins. Benjamini–Hochberg correction was applied and *q <* 0.05 was considered significant.

### 2.8 Pathway Co-localization Testing

Compounds with significant PPI enrichment were tested to determine whether their targets were over-represented in GO biological processes and Reactome pathways previously enriched in the PD biomarker gene set (e.g., “cytokine–cytokine receptor interaction”). Hypergeometric tests were used to evaluate whether the observed overlap between compound targets and pathway genes exceeded random expectation. FDR correction using the Benjamini-Hochberg procedure was applied to account for multiple pathway testing [32].

### 2.9 Composite Severity Index Derivation

A severity index was derived by modeling baseline protein expression against symptom burden. MDS-UPDRS Parts I–IV scores were extracted from AMP-PD clinical datasets and summed to obtain total scores [33], which were then matched to selected baseline features. A linear regression model was trained with five-fold `GroupKFold` cross-validation, using standardized feature coefficients as weights. The resulting severity index represented a weighted sum of baseline features [34, 35].

### 2.10 Longitudinal Progression Modeling

The severity index was evaluated as a predictor of progression using longitudinal samples. Baseline-derived PSI values were projected onto follow-up visits, and a linear mixed-effects model was fitted with total MDS-UPDRS as the dependent variable [36]. Fixed effects included severity index, time (years), and their interaction [37], with random intercepts and slopes specified for each participant. The model was implemented in `statsmodels (MixedLM).`

### 2.11 Statistical Evaluation

Spearman correlations were computed between each selected feature and total MDSUPDRS, with multiple testing correction applied using the Benjamini–Hochberg method. Scatterplots with fitted lines were used to visualize severity index versus MDS-UPDRS, and residuals were assessed to evaluate model fit. Heteroskedasticity was tested using the Breusch–Pagan test.

All analyses were conducted in Python 3.11.4. Core packages included `pandas` (2.2.2), `numpy` (1.26.4), `scikit-learn` (1.4.2), and `statsmodels` (0.14.1). Figures were generated with `matplotlib` (3.8.3).

## 3 Results

### 3.1 Model Comparison and Classification Performance

Proteomic features outperformed other modalities for Parkinson’s disease (PD) classification. Figure 1 summarizes the analytical workflow. Three data configurations were evaluated using the same pipeline: (1) proteomics-only, (2) gene expression–only, and (3) combined proteomics and gene expression. Each configuration was used to train and tune models using a 10-fold `GroupKFold` cross-validation on an 80% training subset and assessed performance on an external PDBP hold-out.

**Fig. 1:**
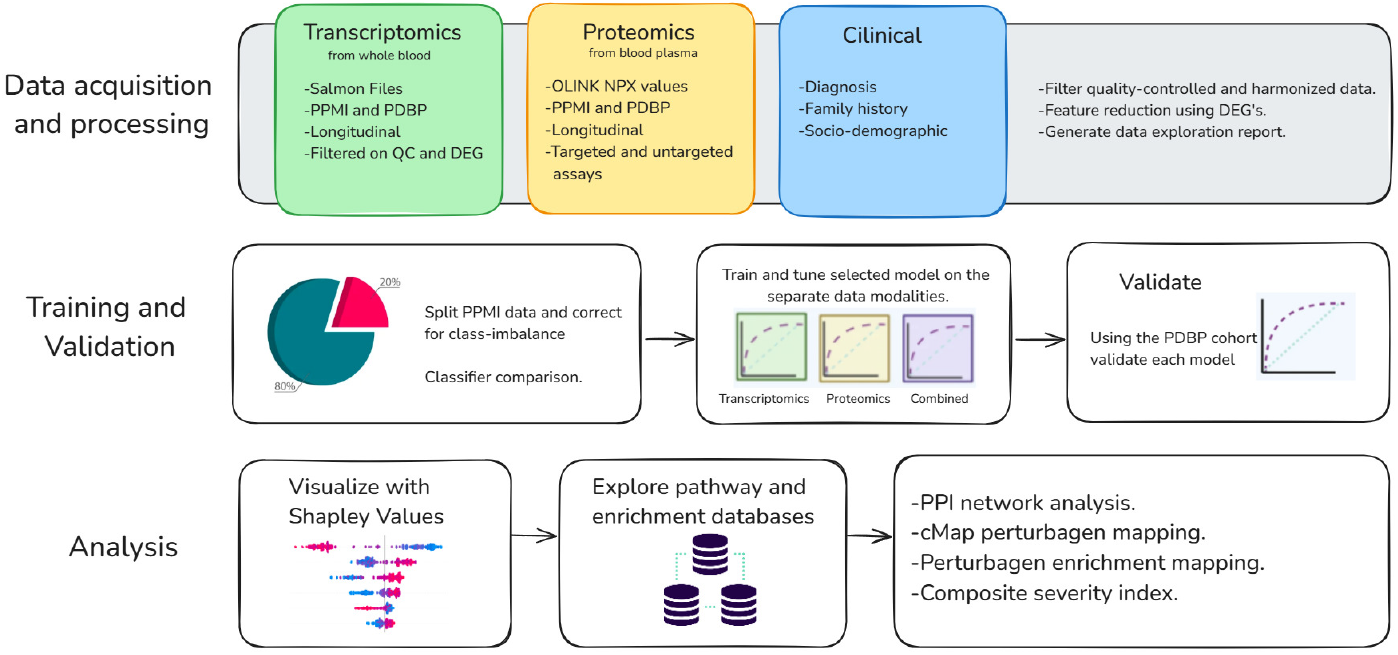
Workflow diagram.

The gene expression–only model performed worst, whereas the proteomics-only model delivered the strongest cross-validated and external performance (Table 1). A paired bootstrap comparison (1,000 resamples) confirmed that the proteomics-only model significantly exceeded the combined model (Wilcoxon signed-rank test, *p* = 1.22 × 10^−103^).

**Table 1.** Model performance across data configurations. ROC-AUC values are reported for cross-validation on the training set and held-out performance on the external validation set. Cross-validation used participant-wise GroupKFold with all preprocessing nested to prevent leakage. Higher AUC indicates better PD-vs-control discrimination; 95% CIs were computed by stratified bootstrap (1,000 resamples).

| Model | Cross-Validation ROC-AUC | Validation ROC-AUC | 95% CI Lower | 95% CI Upper |
| --- | --- | --- | --- | --- |
| Combined | 0.9663 | 0.8825 | 0.8469 | 0.9177 |
| Proteomics only | **0.9710** | **0.8879** | **0.8514** | **0.9236** |
| RNA-seq only | 0.5248 | 0.5869<sup>1</sup> | 0.5507 | 0.6235 |

Source: Results from 10-fold GroupKFold cross-validation and external validation on PDBP cohort.
<sup>1</sup>Proteomics-only model significantly outperforms the Combined model on the validation set (Wilcoxon signed-rank test, $p = 1.22 \times 10^{-103}$ ).

Given its accuracy, parsimony, and cross-cohort robustness, the proteomics-only model was selected for downstream analyses. On the external validation set, this model achieved an ROC–AUC of 0.8879 and an accuracy of 84%. The confusion matrix indicated 36 false negatives and 13 false positives, consistent with high sensitivity and specificity (Figure 2).

**Fig. 2:**
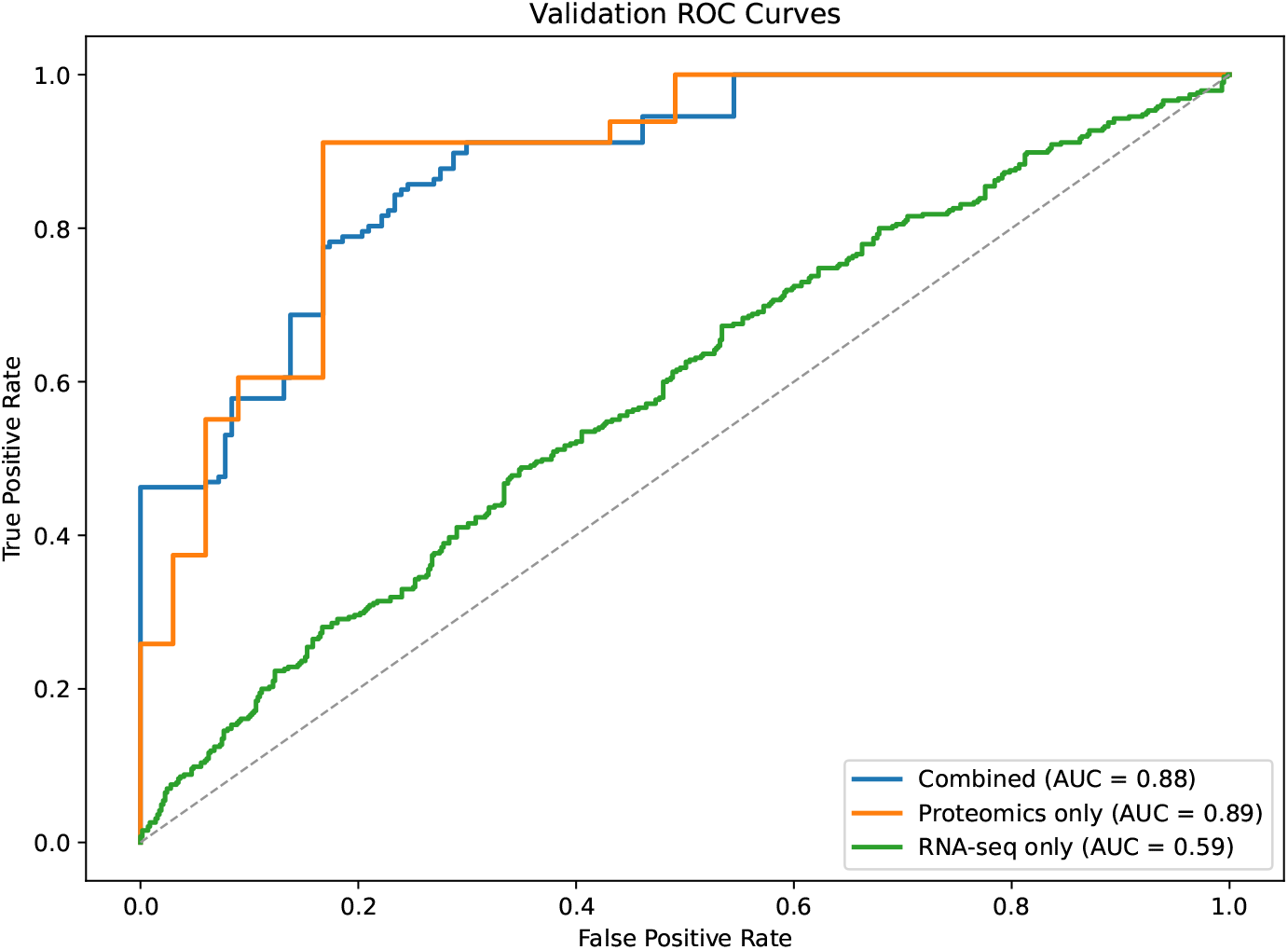
ROC curves for proteomics-only, combined, gene-only models on hold-out set; proteomicsonly highest, gene-only lowest AUC.

Models were optimized with Latin Hypercube Sampling over 50 `RandomizedSearchCV` iterations. The best proteomics-only Random Forest used *n*_*e*_*stimators* = 1074, *max*_*d*_*epth* = 2, *min*_*s*_*amples*_*s*_*plit* = 2, *min*_*s*_*amples*_*l*_*eaf* = 1, and *class*_*w*_*eight* = ‘balanced’. Feature selection retained 32 protein markers that captured the most discriminative signal for PD.

### 3.2 Model Interpretability via SHAP

SHapley Additive exPlanations (SHAP) were used to assess the proteomics-only Random Forest (Figure 3). Because the classifier outputs control-class probability, negative SHAP values indicate features that increase PD probability. `P20711` (*DDC*) showed the largest magnitude contribution (mean SHAP = − 0.063 *±* 0.082), implicating higher levels with increased PD likelihood. `O60760` (*HPGDS*) (0.0121 *±* 0.0518), `Q16853` (*AOC3*) (0.0075 0. *±* 0502), and `P01236` (*PRL*) (− 0.0182 *±* 0.0470) also ranked among the most influential features, with additional moderate effects from `Q7Z5L0` (*VMO1*) (0.0026 *±* 0.0266) and `Q5T2D2` (*TREML2*) (0.0117 *±* 0.0251). Features with the most stable (low-variance) contributions included `Q8NCC3` (*PLA2G15*) (−0.0010 *±* 0.0070), `P21860` (*ERBB3*) (0.0014 *±* 0.0069), and `P35052` (*GPC1*) (−0.0005 *±* 0.0052).

**Fig. 3:**
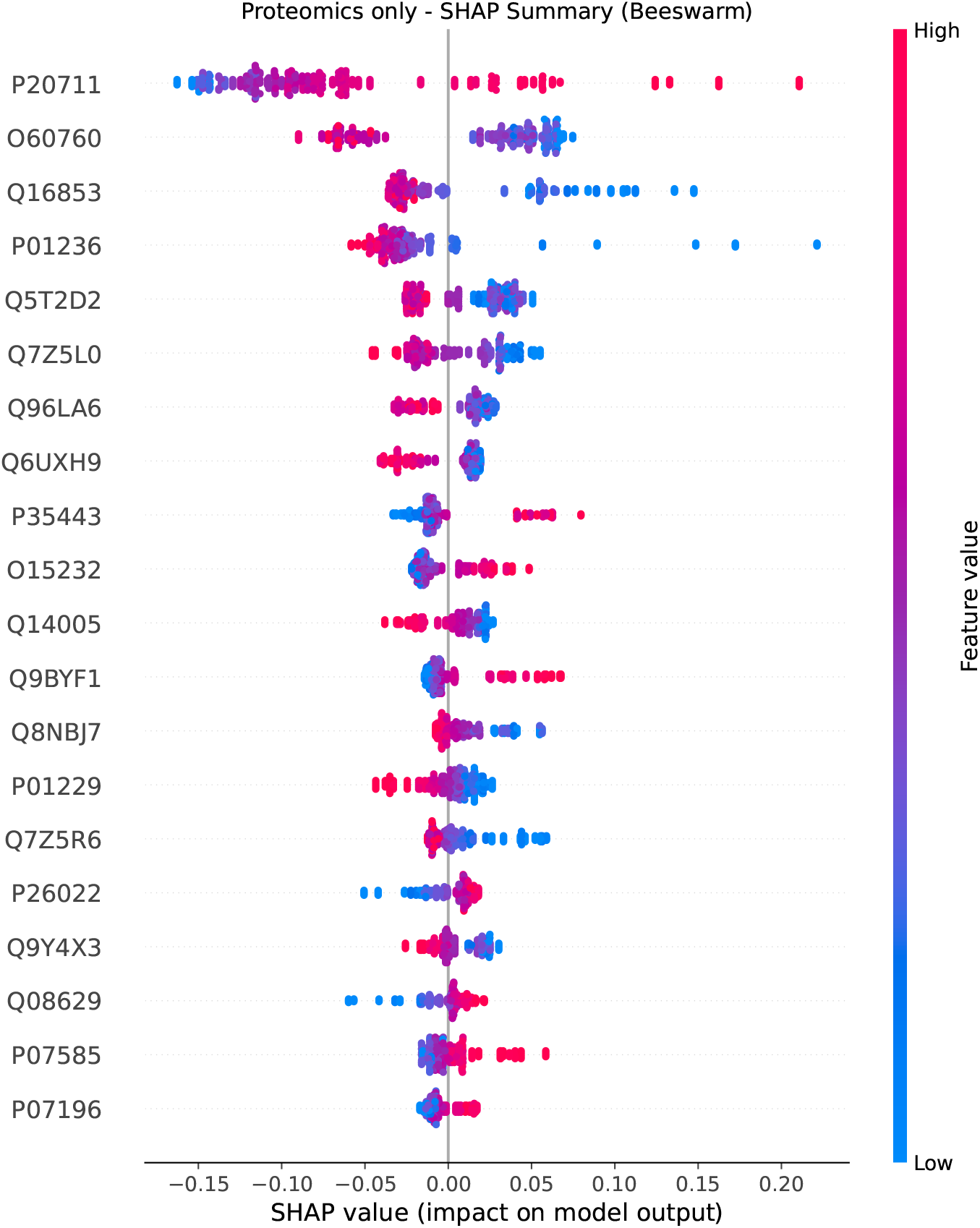
Global feature importance for the proteomics-only classifier. SHAP summary (“beeswarm”) plot for the Random Forest model trained on plasma proteins. Each point is a sample-level SHAP value for a given protein (x-axis: impact on model log-odds). Features are ordered by mean absolute SHAP across participants (top = most influential). Point color encodes the standardized NPX value of the protein in that sample (higher = warmer). Because the classifier outputs control-class probability, negative SHAP values indicate proteins that increase the probability of PD in that sample. Notable contributors include DDC (P20711) and THBS4 (P35443), which have positive associations with PD, and PRL (P01236) and HPGDS (O60760), which have negative associations. Preprocessing (variance filtering, SelectKBest, scaling) and model fitting were nested within 10-fold GroupKFold by participant to prevent leakage; 32 proteins were retained for the final model. See Methods for details.

### 3.3 Gene Ontology Enrichment

Gene Ontology enrichment analysis was conducted on the 32 selected proteins to provide context for their biological functions. Significant over-representation was observed for extracellular-related terms, including “extracellular space” (*p* = 3.0 × 10^−7^), “extracellular region” (*p* = 1.4 × 10^−6^), and “secreted proteins” (*p* = 1.5 × 10^−8^) (Figure 4). Additional enrichments included disulfide bonds (*p* = 3.0 × 10^−9^), glycosylation (*p* = 2.5 × 10^−4^), calcium ion binding (*p* = 9.0 × 10^−4^), and EGF-like domains (*p* = 9.8 × 10^−4^), pointing to signaling programs relevant to neurodegeneration.

**Fig. 4:**
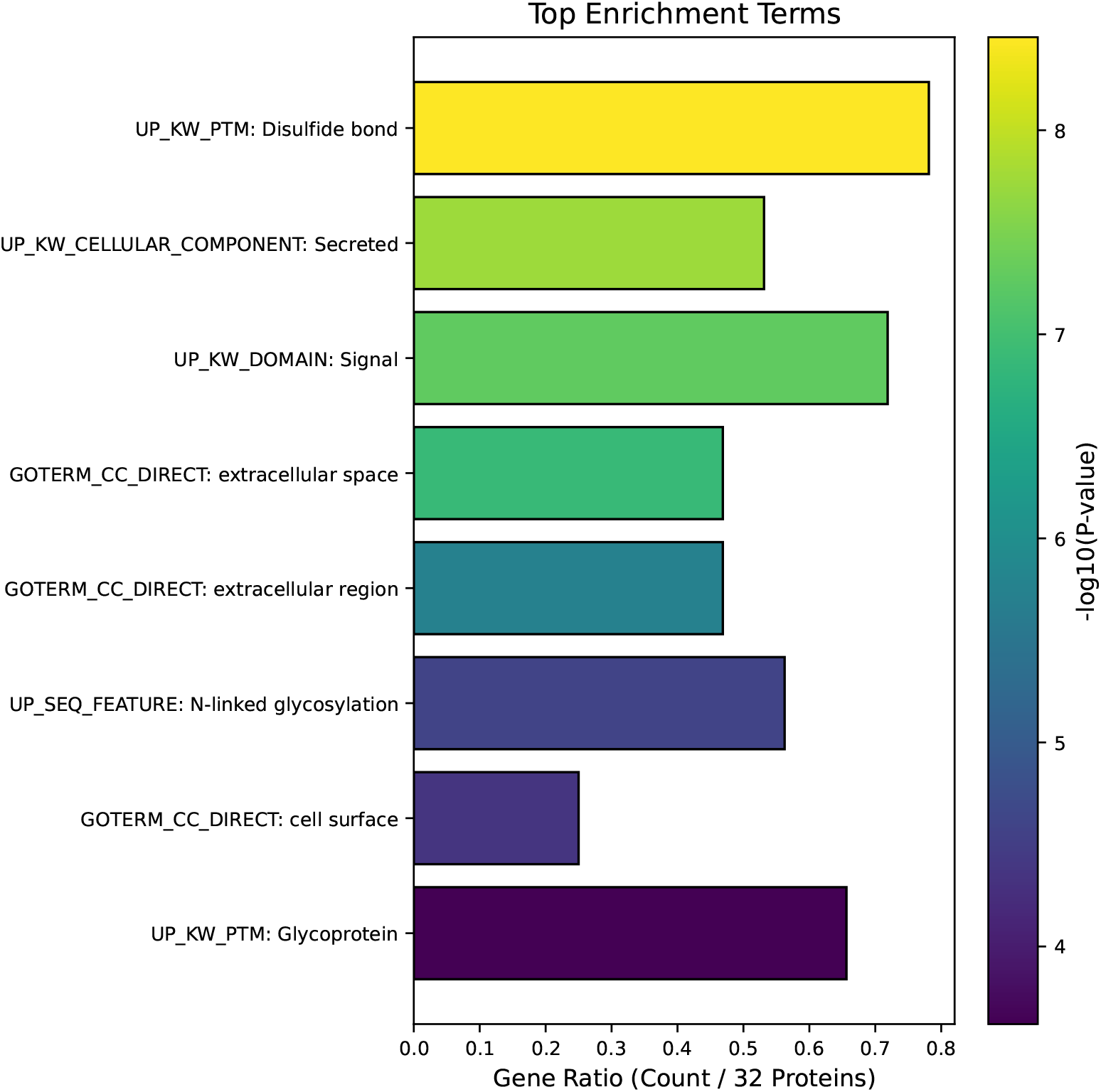
Functional enrichment of the 32 proteins retained by the proteomics classifier. Bar length shows the gene ratio (hits per 32 proteins); color encodes the enrichment significance as − log_10_(*p*). Terms are drawn from UniProt Keywords (UP KW), GO Cellular Component (GOTERM CC DIRECT), and UniProt sequence features. The top enrichments (e.g., *disulfide bond, secreted, signal peptide, extracellular space/region, N-linked glycosylation, cell surface, glycoprotein*) indicate a cystine-rich, glycosylated, extracellular/secreted proteome consistent with circulating plasma biology. Over-representation tests were performed against the assayed protein background with Benjamini–Hochberg FDR control (*q <* 0.05).

Proteins with disulfide bonds (*p* = 3.0 *×*10^−9^), glycosylation (*p* = 2.5 × 10^−4^), calcium ion binding (*p* = 9.0 × 10^−4^), and EGF-like domains (*p* = 9.8 × 10^−4^) were also enriched, suggesting involvement in signaling pathways relevant to neuronal function and neurodegeneration.

### 3.4 Protein–Protein Interaction Network

STRING analysis of the 32 proteins yielded a significantly enriched protein–protein interaction (PPI) network (*p* = 0.0438) with 32 nodes and 10 edges (average degree = 0.625). Functional annotation recapitulated extracellular themes—”extracellular region” (*p* = 2.48 × 10^−6^), “extracellular space” (*p* = 5.40 × 10^−5^)—and highlighted “cytokine–cytokine receptor interaction” (*p* = 0.0020). Central nodes were predominantly secreted, disulfide-bonded, and glycosylated proteins, suggesting roles in immune signaling and inflammation. Enrichment of proteoglycans (*p* = 0.0151) and heparan sulfate–related proteins (*p* = 0.0430) was also observed, implicating extracellular matrix pathways.

### 3.5 Drug Perturbation Mapping

Differential expression analysis confirmed that all 32 selected proteins were significantly altered in PD versus controls. Overexpressed proteins included `P20711` (*DDC*) (fold change = 2.17), `P35443` (*THBS4*) (2.08), and `P01130` (*LDLR*) (1.47), while underexpressed proteins included `P01236` (*PRL*) (− 5.02), `Q96N03` (*VSTM2L*) (− 6.12), and `Q9BYF1` (*ACE2*) (− 2.00), consistent with dysregulated immune and metabolic processes.

This analysis step is hypothesis-generating; results depend on the CLUE build and filters used. Connectivity Map (CMap) analysis identified three compounds with strongly negative connectivity scores, suggesting potential reversal of PD-associated expression patterns (Appendix Table A). Each candidate’s validated targets were projected onto a curated Parkinson’s disease protein–protein interaction (PPI) network. Among these, only the top-ranked perturbagen, SAL-1 (alloxazine/isoalloxazine), was retained. Annotated in CMap as an adenosine receptor antagonist targeting ADORA2A/ADORA2B and profiled in L1000 (Broad Repurposing Hub ID BRDK40213712). SAL-1 overlapped with PD-associated nodes. GO and Reactome analyses of SAL-1 targets revealed 45 enriched pathways at *q* ≤ 0.05, including circadian rhythm regulation, purinergic signaling, and glutamate/GABA transmission, supporting SAL-1 as a mechanistically plausible candidate for follow-up.

### 3.6 Protein Associations with Clinical Severity

Plasma protein levels were correlated with total MDS-UPDRS scores. `P20711` (*DDC*) showed the strongest positive association (*ρ* = 0.61, *q* = 5.9 × 10^−11^), followed by `P07196` (*NEFL*) (*ρ* = 0.44, *q* = 2.3 × 10^−5^). Prolactin (`P01236`, *PRL*; *ρ* = −0.43) and `O60760` (*HPGDS* ; *ρ* = −0.30) were negatively correlated (Figure 5).

**Fig. 5:**
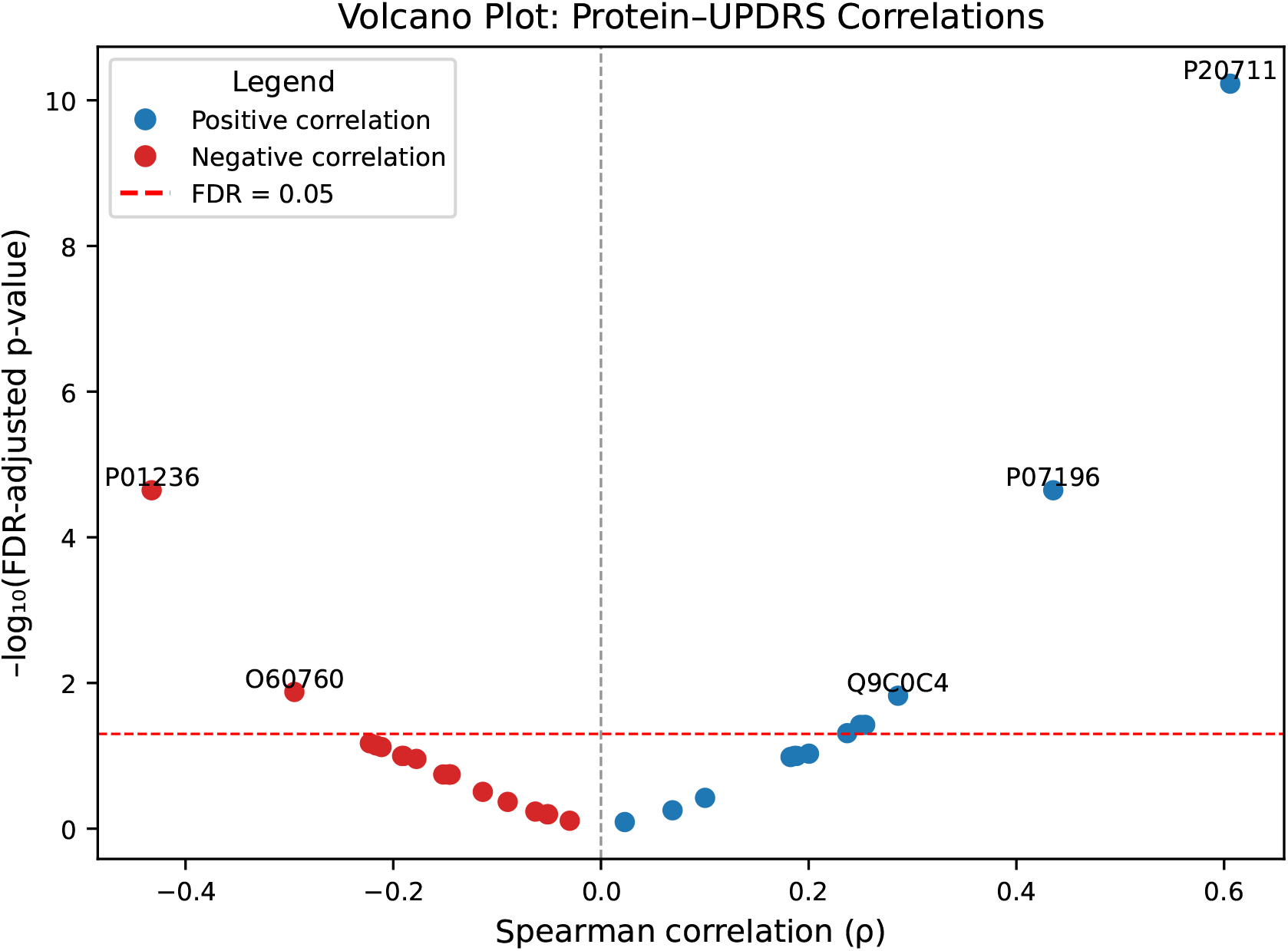
Plasma proteins correlated with clinical severity (MDS-UPDRS total). Volcano plot of protein–severity associations at baseline (Spearman *ρ* on the x-axis; − log_10_(FDR-adjusted *p*) on the y-axis). Associations were computed within PD participants (baseline draw; multiple testing controlled by Benjamini–Hochberg). Proteins to the right are positively correlated with greater motor or non-motor burden; to the left are negatively correlated (putative protective/compensatory patterns). Representative signals are annotated: DDC (P20711) and NEFL (P07196) show positive correlations, whereas PRL (P01236) and HPGDS (O60760) are negatively correlated. Full statistics are provided in Supplementary Tables; analysis choices and quality controls are described in Methods.

### 3.7 Derivation of Proteomic Severity Index (PSI)

A Proteomic Severity Index (PSI) was constructed as a weighted linear combination of the 32 proteins. Among 111 participants with baseline data, PSI explained 28.2% of the variance in total MDS-UPDRS (mean cross-validated *R*^2^ = 0.282). The largest positive weights were assigned to `P20711` (*DDC*) (+9.70) and `P35443` (*THBS4*) (+7.66), whereas `Q16853` (*AOC3*) (− 5.73), `P01229` (*LHB*) (− 4.80), and `P01236` (*PRL*) (− 3.74) carried negative weights, indicating inverse associations with symptom burden.

### 3.8 Longitudinal prediction of disease progression

A total of 745 longitudinal observations from 235 participants were modeled using linear mixed-effects. Baseline PSI independently predicted total MDS-UPDRS (sum of Parts I–IV; *β* = 0.277, *p <* 0.0001), and time was also positively associated (*β* = 1.80, *p <* 0.0001). The PSI × time interaction was negative and significant (*β* = − 0.043, *p* = 0.027), suggesting that individuals with higher baseline PSI experienced slightly slower symptom progression over follow-up.

## 4 Discussion

Our findings demonstrate that proteomic features can outperform transcriptomic and multimodal features in distinguishing Parkinson’s disease (PD) from healthy controls. As the functional endpoints of gene expression, proteins may more accurately reflect ongoing biological processes. Because proteins are the effectors of cellular function, proteomic measurements more directly reflect disease activity and can reveal changes not captured by RNA-seq. Additionally, proteomic profiles exhibited greater cross-cohort consistency, underscoring their translational potential for PD detection and patient stratification.

Strong enrichment for extracellular and secreted proteins was observed, indicating that Parkinson’s disease–related changes extend beyond intracellular signaling into circulating compartments accessible in plasma. Many of these proteins mediate intercellular communication, immune modulation, and extracellular matrix (ECM) remodeling, processes increasingly implicated in neurodegeneration [38]. Enrichment for post-translational features such as disulfide bonds and glycosylation supports specialized extracellular functions, including stabilization, trafficking, and receptor engagement [39]. The protein–protein interaction network contained more edges than expected by chance, which is consistent with coordinated activity among circulating factors. Together, these observations support a model in which PD involves measurable disruption of the plasma proteome, particularly among extracellular signaling and structural proteins that could serve as biomarkers or therapeutic entry points.

The prominence of EGF-like domains suggests additional ligand–receptor pathways that remain underexplored in PD [40]. Concurrent enrichment for cytokine–cytokine receptor interactions reinforces the role of neuroinflammation, including microglial activation and cytokine release, in dopaminergic neuron loss [41]. Calcium-binding terms and synaptic pathway annotations further point to perturbed neurotransmission, consistent with reports of altered synaptic proteins in brain-derived extracellular vesicles from PD patients [42]. Reactome analysis converged on `GPC1`, `DCN`, `THBS4`, `MATN3`, `ERBB3`, and `PDGFC`, implicating ECM proteoglycan remodeling, glycosaminoglycan metabolism, and receptor tyrosine kinase signaling. Although typically linked to tissue remodeling and fibrosis, these pathways may impair neurotrophic signaling and amplify inflammatory tone in PD [43], and they merit targeted investigation.

Several of these blood-based proteins have been repeatedly linked to PD risk or progression in prior literature, including neurofilament light chain (NfL), glial fibrillary acidic protein (GFAP), total/oligomeric *α*-synuclein, DJ-1 (PARK7), and proinflammatory cytokines (e.g., IL-6, TNF-*α*).[44–46]. Large proteomic cohorts have also highlighted plasma proteins associated with incident or prodromal PD [47]. Within our panel, aromatic L-amino acid decarboxylase (AADC/DDC), a dopamine-synthesis enzyme, has recently emerged as a fluid biomarker measurable in plasma and linked to Lewy body disorders and parkinsonian syndromes.[48, 49]. By contrast, circulating prolactin (PRL) shows mixed evidence in PD and is sensitive to dopaminergic therapy effects,[50, 51] while hematopoietic prostaglandin D synthase (HPGDS) has mechanistic support in neuroinflammation but limited validation as a PD blood biomarker.[52]. Our findings both recapitulate emerging plasma signals (e.g., DDC) and nominate less-explored extracellular proteins that align with immune/ECM pathways for further validation.

### Potential therapeutics

Our *in silico* perturbation analysis identified small molecules with strongly negative connectivity to the PD proteomic signature. SAL-1 (alloxazine/isoalloxazine) emerged as a notable candidate, consistent with reported adenosine receptor antagonism at A_2B_ and potential A_2A_ activity [53]. While dopaminergic therapies remain standard of care, they do not slow disease progression and can cause dyskinesia [54]. Modulating adenosine signaling, particularly A_2A_ pathways, may improve motor symptoms and reduce dyskinesia, though SAL-1 has not been studied in PD. These results are hypothesis-generating and require experimental validation.

Fostamatinib, a SYK inhibitor, and homoharringtonine (HHT), a protein synthesis inhibitor, also showed negative connectivity. Prior integrative analyses have implicated fostamatinib through interactions with targets such as `DCLK1`, `KCNJ6`, and `SNCA` [55]. HHT reduced amyloid burden and neuroinflammation in Alzheimer’s disease models via STAT3 signaling [56], which suggests relevance to shared inflammatory mechanisms in PD. Conversely, compounds with positive connectivity, including the proteasome inhibitor MG-132, are known to exacerbate proteostatic stress and promote *α*-synuclein aggregation [57]. PKC activators such as prostratin and phorbol-12-myristate-13-acetate may similarly amplify maladaptive kinase cascades [55, 58]. These contrasts highlight therapeutic strategies that restore protein homeostasis, dampen inflammation, and normalize dysregulated receptor signaling.

### Proteomic Severity Index

Several proteins showed directionally consistent associations with total MDS-UPDRS. `P20711` (*DDC*), an enzyme in dopamine biosynthesis, was the top positive correlate and the strongest contributor to our severity index, aligning with biological expectations. Prolactin (`P01236`, *PRL*) and hematopoietic prostaglandin D synthase (`O60760`, *HPGDS*) correlated negatively with symptom burden, which may indicate protective or compensatory responses early in disease.

The Proteomic Severity Index (PSI) discriminated moderate-to-severe cases with an AUROC of 0.964 and explained 28.2% of the variance in total MDS-UPDRS, which is notable in a multifactorial disorder [59, 60]. In longitudinal models, PSI and time each showed independent effects on severity. The negative PSI-by-time interaction suggests that individuals with high baseline PSI exhibited slightly slower subsequent worsening. This pattern could reflect regression toward the mean, therapeutic exposure, ceiling effects, or underlying biology. Prospective validation is needed to distinguish among these explanations.

Overall, PSI emerges as a reproducible, blood-based index of motor symptom burden with potential utility for patient stratification and progression monitoring, including trial enrichment and endpoint refinement.

### Limitations

This study uses multi-site PPMI and PDBP datasets, which introduce variability from demographics, sampling protocols, and batch effects [61]. Such heterogeneity can dilute effect sizes and complicate generalization across settings [62]. Peripheral blood biomarkers provide practical accessibility but may incompletely capture central nervous system processes central to PD pathogenesis. The proteomics sample size and feature count constrain model complexity. Although rigorous cross-validation, external testing, and conservative feature selection were employed, data-driven selection may still introduce overfitting or selection bias. Finally, the primary analysis is cross-sectional, which limits causal inference about temporal dynamics.

### Biological validation

Computational robustness does not replace experimental proof. Independent cohorts, targeted protein assays, and mechanistic work in cellular and animal models are necessary to establish causality. Candidate therapeutics from the connectivity analysis require preclinical testing for target engagement, efficacy, and safety before any clinical consideration.

### Future work

Future studies should validate and extend the drug-repurposing signals and pathway-level hypotheses reported here. SAL-1 and fostamatinib merit particular attention, given potential links to receptor tyrosine kinases, ECM remodeling, and immune signaling. The observed extracellular signatures, including proteoglycan and glycosaminoglycan pathways, support the rationale for multi-tissue profiling in cerebrospinal fluid, brain tissue, and induced pluripotent stem cell–derived neuronal models. Integrating spatial transcriptomics and phosphoproteomics could clarify how extracellular changes intersect with neuronal signaling at cellular resolution.

Clinically, PSI is a scalable, minimally invasive tool that warrants evaluation for predicting conversion from prodromal to clinical PD and for responsiveness to pharmacologic interventions. Embedding PSI into adaptive trial designs could enable biomarker-guided enrichment and more sensitive endpoints [63]. Defining molecular subtypes may further align therapeutic mechanisms with specific patient groups [64]. Extending this framework to other neurodegenerative diseases may reveal shared or distinct biomarker programs that inform cross-disease repurposing and systems-level understanding.

## 5 Conclusion

Proteomics-based models distinguished PD from healthy controls, revealed extracellular and immune-linked biology, and yielded a blood-based severity index with prognostic value. These findings highlight the translational promise of plasma proteomics for PD. Focused experimental validation and collaborative clinical studies are the next steps toward therapeutic development and biomarker deployment.

## Data Availability

All data analyzed in this study were obtained from the Accelerating Medicines Partnership Parkinsons Disease (AMP PD) Knowledge Platform (https://amp-pd.org), which includes data from the PPMI and PDBP cohorts. Access to these de-identified datasets requires registration, data use agreements, and approval by AMP PD. Derived data supporting the findings of this study are available from the corresponding author upon reasonable request.

https://github.com/NMinster/pd-multiomics-analysis

## 6 Availability of data and materials

Data statement here. Code statement here.

## Abbreviations

AMP-PD: Accelerating Medicines Partnership Parkinson’s Disease
AUC: Area Under the Curve
CLUE: Connectivity Map User Environment
CMap: Connectivity Map
CNS: Central Nervous System
ECM: Extracellular Matrix
EV: Extracellular Vesicle
FDR: False Discovery Rate
GO: Gene Ontology
LHS: Latin Hypercube Sampling
MAO-B: Monoamine Oxidase B
MDS-UPDRS: Movement Disorder Society Unified Parkinson’s Disease Rating Scale
MRI: Magnetic Resonance Imaging
NGS: Next-Generation Sequencing
NPX: Normalized Protein Expression
PD: Parkinson’s Disease
PDBP: Parkinson’s Disease Biomarkers Program
PEA: Proximity Extension Assay
PPI: Protein–Protein Interaction
PSI: Proteomic Severity Index
13 q: Adjusted p-value (FDR-corrected)
RNA-seq: RNA Sequencing
ROC: Receiver Operating Characteristic
SHAP: SHapley Additive exPlanations
STRING: Search Tool for the Retrieval of Interacting Genes/Proteins
SPECT: Single Photon Emission Computed Tomography
TPM: Transcripts Per Million

## Declarations

### Ethics approval and consent to participate

A secondary analysis was conducted using de-identified, controlled-access individual-level data from the AMP-PD Knowledge Platform (PPMI IR3 and PDBP 2023 releases). Access required a Data Use Agreement (DUA) authorized by George Mason University. AMP-PD does not provide direct identifiers or any re-identification key to investigators, and the DUA prohibits any attempt to re-identify participants. All AMP-PD data-handling requirements and George Mason University research data policies were followed.

The IRB at George Mason University determined that IRB ID: STUDY00000792 is not research involving human subjects as defined by DHHS and FDA regulations.

The source cohorts (PPMI and PDBP) obtained ethics approval at participating institutions and collected written informed consent from all participants before data collection and de-identification. No contact or interaction with participants occurred, and no interventions were performed. **Consent for publication**

Not applicable. This study includes no individual person data (images or identifiable details).

### Availability of data and materials

Data used in this study were obtained from the Accelerating Medicines Partnership Parkinson’s Disease (AMP-PD) Knowledge Platform (https://amp-pd.org). Access to AMP-PD requires registration and approval by the respective data access committees.

All code used for preprocessing, modeling, and statistical analysis is openly available at [https://github.com/NMinster/pd-multiomics-analysis]. Processed data files sufficient to reproduce the figures and tables in this manuscript are provided in the same repository, subject to the AMP-PD and PDBP data use agreements.

### Competing interests

The authors declare that they have no competing interests.

### Funding

This work was supported by the National Science Foundation Research Traineeship (NSF-NRT) program and the George Mason University Dissertation Completion Grant. The authors and their institution did not receive payment or services from a third party for any other aspect of the submitted work.

### Authors’ contributions

Author 1 conceptualized the study, performed data analysis, interpreted results, and wrote the manuscript. Author 2 supervised the project, contributed to the study design and interpretation, and revised the manuscript for critical content. Both authors read and approved the final manuscript.

## Acknowledgements

Data used in the preparation of this article were obtained from the Accelerating Medicines Partnership^®^ (AMP^®^) Parkinson’s Disease (AMP^®^ PD) Knowledge Platform. For up-to-date information on the study, visit https://www.amp-pd.org.

The AMP^®^ PD program is a public–private partnership managed by the Foundation for the National Institutes of Health and funded by the National Institute of Neurological Disorders and Stroke (NINDS) in partnership with the Aligning Science Across Parkinson’s (ASAP) initiative; Celgene Corporation, a subsidiary of BristolMyers Squibb Company; GlaxoSmithKline plc (GSK); The Michael J. Fox Foundation for Parkinson’s Research; Pfizer Inc.; AbbVie Inc.; Sanofi US Services Inc.; and Verily Life Sciences.

*ACCELERATING MEDICINES PARTNERSHIP and AMP are registered service marks of the U*.*S. Department of Health and Human Services*.

PPMI is sponsored by The Michael J. Fox Foundation for Parkinson’s Research and supported by a consortium of scientific partners. For up-to-date information on the study, visit https://www.ppmi-info.org.

The Parkinson’s Disease Biomarker Program (PDBP) consortium is supported by the National Institute of Neurological Disorders and Stroke (NINDS) at the National Institutes of Health. A full list of PDBP investigators can be found at https://pdbp.ninds.nih.gov/policy. The PDBP investigators have not participated in reviewing the data analysis or content of the manuscript.

## Authors’ information

Not applicable.

## Appendix A cMap Results

**Table A1.**
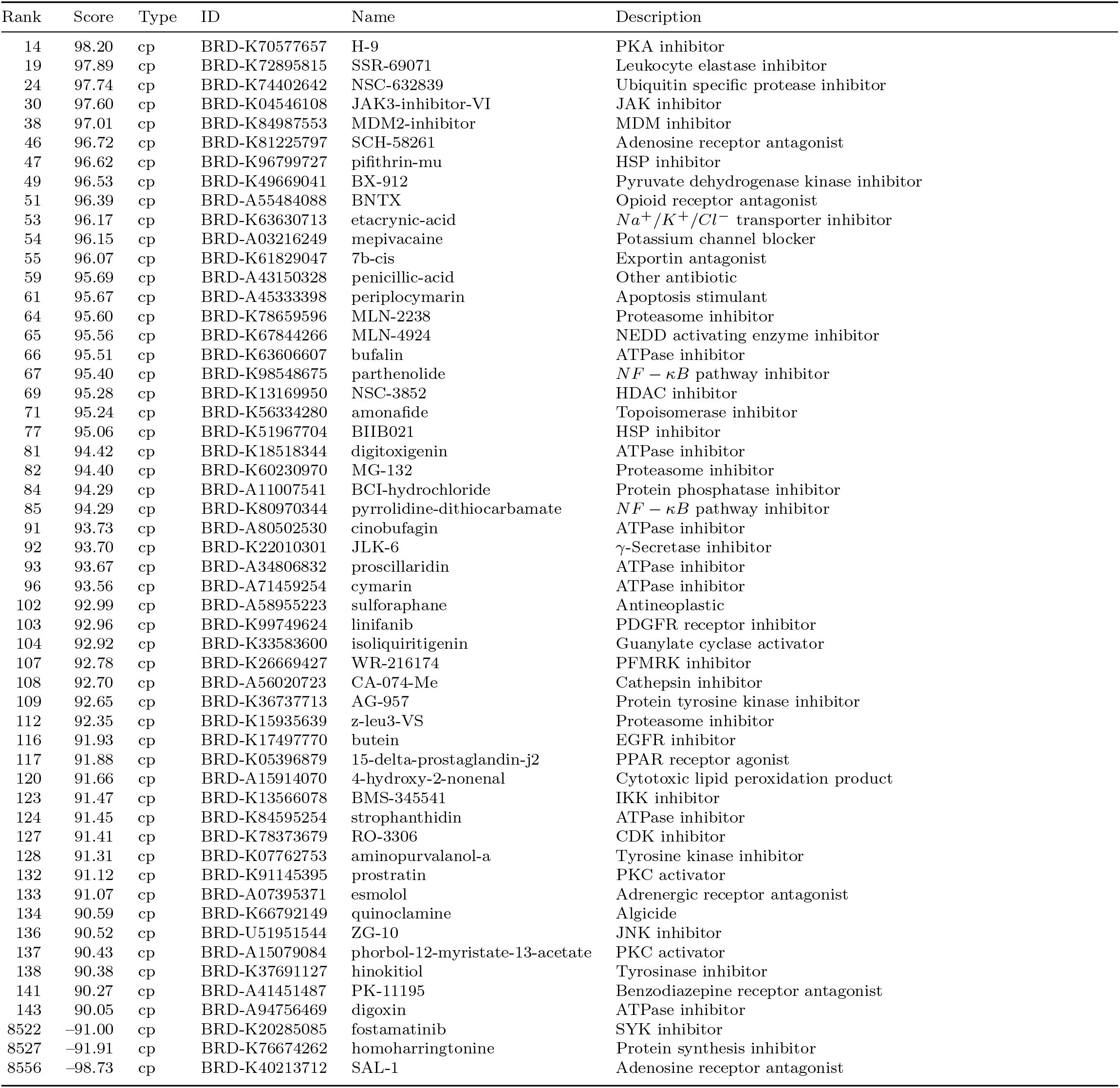
High-scoring compounds from screening (sideways layout)

## Notes

### Competing Interest Statement

The authors have declared no competing interest.

### Author Declarations

Institutional Review Board of George Mason University waived ethical approval for this work.

### Summary of Updates

1. Added co-author 2. Third person. Removed the use of we. 3. Updated abstract and background with the value of early diagnosis. 4. Explicitly stated three biomarker uses (i) diagnostic classification, (ii) severity monitoring via PSI, (iii) hypothesis generation for therapeutics. 5. Clarified that predictors are RNA-seq + Olink features for binary PD vs healthy control. 6. Hyperparameter search rationale added (Latin Hypercube sampling) with citation. 7. Added SHAP package/version 8. Consistent nomenclature, e.g. MDS-UPDRS used throughout, replaced inconsistent use of UPDRS and MDS-UPDRS 9. CMap results framed as hypothesis-generating and build dependency noted. 10. Figure captions expanded 11. Protein IDs annotated with gene symbols 12. Table 1 caption expanded 13. New paragraph linking to prior blood biomarkers.

